# Postoperative Surveillance Adherence in Colorectal Cancer Patients at Urban Medical Center: is it Adequate?

**DOI:** 10.1101/2024.05.23.24307790

**Authors:** Ahmad Alnasarat, Talin R. Darian, Awni Shahait, Mohanad Baldawi, Gamal Mostafa

## Abstract

**Aim:** To examine the adherence rate to surveillance guidelines after curative resection of colorectal cancer (CRC) and the impact of demographic factors.

**Methods:** Data was collected retrospectively including demographics, stage at diagnosis, and adherence to surveillance guidelines as recommended by the US Multi-Society Task Force (USMSTF) guidelines, for colorectal cancer (CRC) patients who underwent curative surgical resection between 2005 and 2014 in a tertiary academic medical center.

**Results:** A total of 124 patients were included (Male /female, 56.5%/ 43.5%), African American 109 (87.9%), and 70 patients (56.5%) had Medicare/Medicaid insurance. Overall, Appropriate clinical evaluation twice per year for 3 years following surgery was completed in 78 (63%) of patients. A total of 56 (45%) had carcinoembryonic antigen (CEA) levels checked twice a year for 3 years. Surveillance colonoscopy 1 year postoperatively occurred in 64 (51.6%), and 37 (29.8%) had a second colonoscopy 3 years postoperatively. Abdomen/pelvis CT scan was obtained in 90 (72.5%) at 1 year post-operatively. In the entire cohort, strict adherence to post-recession surveillance only occurred in 46 (37.1%). There was no correlation between adherence to surveillance and gender (*p*=0.184), race (*p*=0.118), or insurance type (*p*=0.51).

**Conclusion:** Adherence to surveillance after curative resection of CRC was inadequate regardless of socioeconomic, medical insurance, or race. Measures should be taken to identify barriers and improve compliance with guidelines.

**What is already known on this topic?:** Effective surveillance after colorectal cancer surgery is essential, yet many patients fail to follow recommended guidelines. Existing research highlights disparities in CRC outcomes based on race, socioeconomic status, and insurance coverage, affecting early detection and treatment.

**What this study adds:** Our study reveals a 37.1% adherence rate to surveillance guidelines among CRC patients in an urban medical center, unaffected by gender, race, or insurance status. This challenges previous assumptions about demographic disparities in post-resection care.

**How this study might affect research, practice, or policy:** The study highlights the need for interventions to improve adherence rate to CRC surveillance. Future research, clinical practices, and policy initiatives should focus on overcoming barriers to ensure equitable and effective follow-up care.

## Introduction

Colorectal cancer (CRC) is the second most common cause of cancer-related mortality in the United States^1^. In 2023, an estimated 153,020 individuals were diagnosed with CRC, with approximately 52,550 deaths attributed to the disease^1^. CRC-related mortality has declined over the past couple of decades^1^. This is primarily due to improved early detection with screening colonoscopy, surgical advances, and also new targeted chemotherapy^1^. However, there is documented disparity in the early detection rate and mortality of CRC that is related to ethnicity, socioeconomic and insurance status^2–4^. In general, insurance and socioeconomic status as well as race, were found to have a significant impact on screening, early detection, and mortality of CRC^4^. This could be explained by the impact of these factors on access to medical care and treatment^5^.

The optimal treatment of non-metastatic CRC is curative resection followed by appropriate surveillance to detect early recurrence. There are several established surveillance guidelines following a curative surgical resection of CRC. These guidelines are defined by the United States Multi-Society Task Force on Colorectal Cancer (USMSTF)^6,7^, this is a group of leading gastroenterologists from the American College of Gastroenterology, the American Gastroenterological Association, and the American Society for Gastrointestinal Endoscopy^8^.

We conducted a retrospective review aiming to examine the adherence rate to post-resection surveillance guidelines of CRC cases in urban tertiary medical centers to determine the potential impact of demographic factors that are known to affect screening, on the surveillance rate of these cases.

## Methods

We conducted a retrospective review of patients who underwent curative resection of CRC at two urban teaching medical centers over 9 years (January 2005, through December 2014). For the purpose of the study, curative surgical resection was defined as achieving adequate clear surgical margins with lymph nodes harvesting according to established operative oncological principles in patients without evidence of systemic disease. Exclusion criteria include patients who did not undergo any treatment, patients who received only chemotherapy or radiation for palliative purposes, and patients who underwent palliative surgical resection. Patients who underwent neoadjuvant chemotherapy followed by curative resection were included.

Surveillance procedures were defined according to the guidelines established by the USMSTF^8^. This included a protocol of office visits every 3-6 months for the initial 3 years, serum Carcinoembryonic Antigen (CEA) level monitoring at each follow-up during the first 3 years, annual computed tomography (CT) scans of the chest, abdomen, and pelvis for a minimum of the initial 3 years, and a strategically timed repeat colonoscopy 1-year post-operation, followed by subsequent colonoscopies every 3-5 years contingent upon findings from the initial 1-year colonoscopy.

Data collection included patient demographics, oncological data, operative details, and postoperative surveillance. Data was presented as numbers, percentages, and ratios as appropriate. Numerical variables were analyzed with *t*-test or ANOVA and categorical variables with Pearson Chi-squared or Fisher exact test as appropriate. Regression analysis was applied to determine independent predictors of outcome. In all analyses, a p-value ≤ 0,05 was considered statistically significant. All statistical analyses were conducted utilizing SPSS (IBM Corp. Released 2017. IBM SPSS Statistics for Windows, Version 25.0). Ethical approval for this study was obtained from the Institutional Review Board of Wayne State University in Detroit, Michigan.

## Results

The initial search, employing the keywords “colon cancer” and “rectal cancer,” yielded a cohort of 698 patients. Out of these patients, 574 were excluded according to our exclusion criteria resulting in a final cohort of 124 patients who underwent curative surgical resection ().

**Figure 1.**
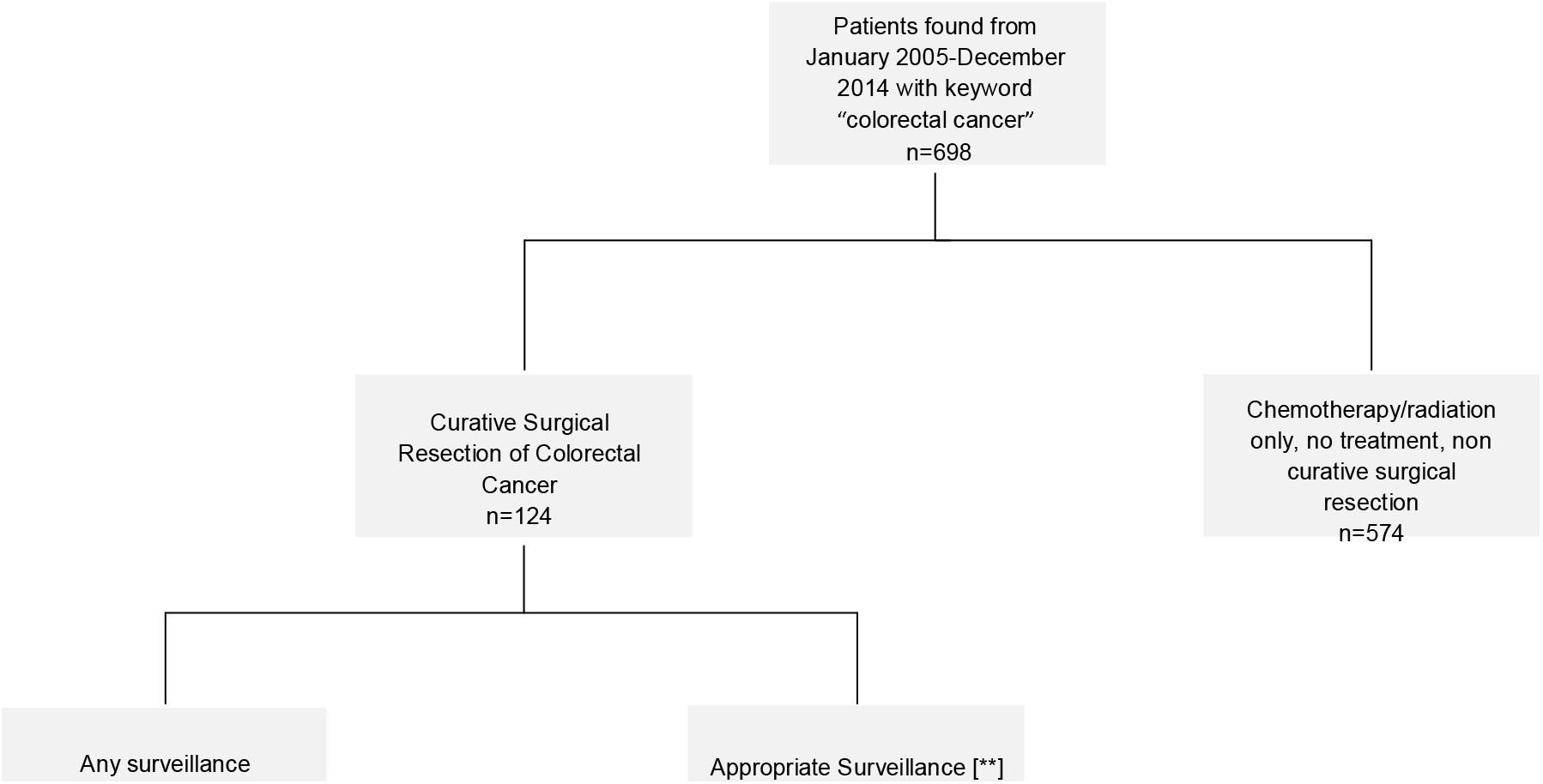
Model for Patient Selection of Curative Surgical Resection. **\*\*=per USMSTF guidelines**

The demographic features, insurance status, and cancer stage at the time of diagnosis are shown in ***Table 1*. Relation between Patient Characteristics and CRC stage at Time of Diagnosis** As shown, neither gender (*p*=0.746, degree of freedom (df)=4), race (*p*=0.122, df=8), nor insurance status (*p*=0.472, df=8) emerged as a predictor of the stage of cancer at the time of diagnosis.

**Table 1.**
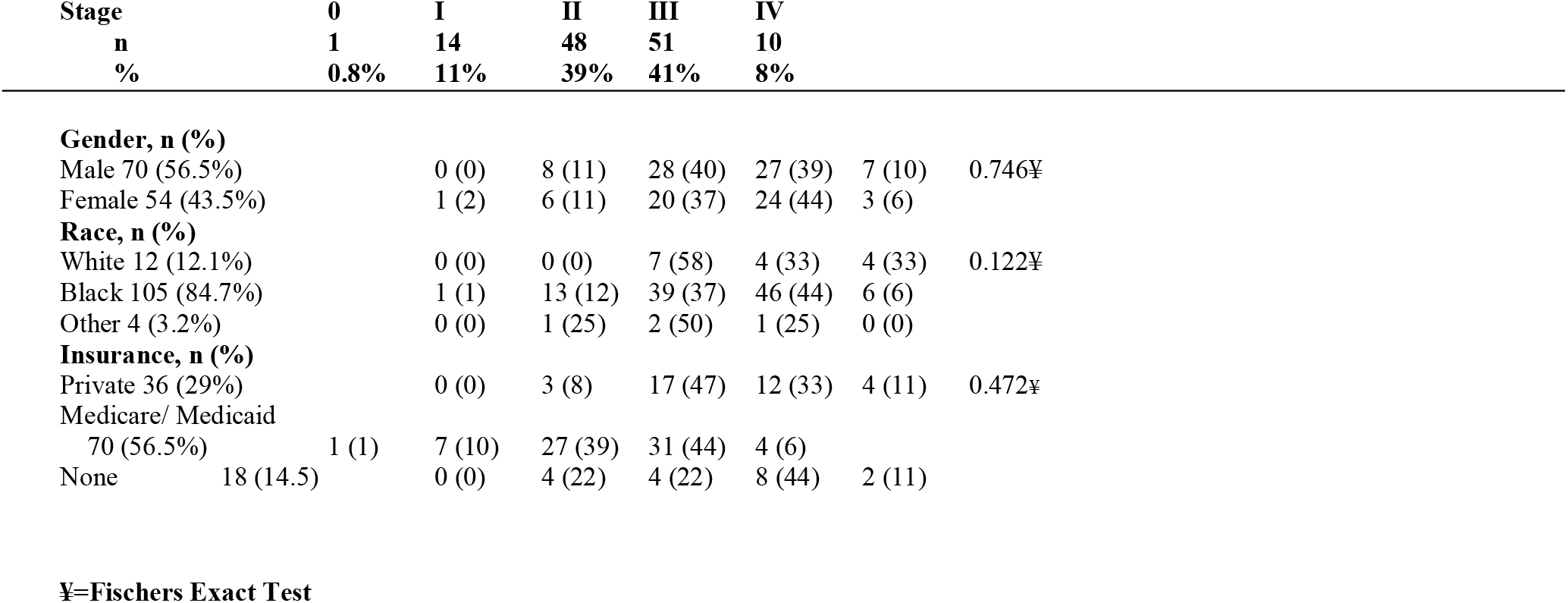
Relation between Patient Characteristics and CRC stage at Time of Diagnosis.

The relationship between demographics and adherence to postoperative surveillance in all patients is shown in **Table 2. Patients’ Characteristics Comparing Any Surveillance vs. Appropriate Surveillance. \*\*** Overall, 105 patients (86%) underwent some form of surveillance, while only 46 (37.7%) adhered to the “appropriate surveillance” criteria outlined by the USMSTF. Overall, Female patients received a higher percentage of some element to surveillance 49 (91%) compared to males. In addition, 21 (39%) of female patients adhered to the standard of strict surveillance compared to males 25 (36%). The gender difference (*p*=0.184, df=1) in receiving any measure of surveillance or adherence to strict surveillance criteria did not reach statistical significance. Neither was there any statistically significant difference in adherence to surveillance based on race (*p*=0.118, df=2), insurance status (*p*=0.513, df=2) or stage at time of diagnosis. (**Table 2. Patients’ Characteristics Comparing Any Surveillance vs. Appropriate Surveillance. \*\***.

**Table 2.** Patients’ Characteristics Comparing Any Surveillance vs. Appropriate Surveillance. **.

|  | Any<br>Surveillance | p-value | Appropriate<br>Surveillance | p-value |
| --- | --- | --- | --- | --- |
| **Gender, n (%)** |  |  |  |  |
| Male | 56 (80) | 0.184 | 25 (36) | 0.810 |
| Female | 49 (91) |  | 21 (39) |  |
| **Race, n (%)** |  |  |  |  |
| White | 12 (100) | 0.118 | 7 (48) | 0.201¥ |
| Black | 92 (88) |  | 39 (37) |  |
| Other | 2 (50) |  | 0 (0) |  |
| **Insurance, n (%)** |  |  |  |  |
| Private | 32 (89) | 0.513 | 15 (42) | 0.811 |
| Medicare/<br>Medicaid | 59 (84) |  | 24 (34) |  |
| None | 14 (78) |  | 7 (39) |  |
| **Stage, n (%)** |  |  |  |  |
| 0 | 1 (100) | 0.129 | 0 (0) | 0.819¥ |
| I | 9 (64) |  | 5 (36) |  |
| II | 42 (88) |  | 16 (33) |  |
| III | 45 (88) |  | 22 (43) |  |
| IV | 8 (80) |  | 3 (30) |  |

\*=significant, ¥= Fischers Exact Test, \*\*=per USMSTF guidelines

Sub-analysis was performed to determine the adherence to specific surveillance measures individually. A total of 78 (62.9%) patients underwent office clinical evaluation by their physician twice a year for 3 years post-surgery. Colonoscopy was performed 1-year post-operatively in 64 (51.6%) patients and only 37 (29.8%) of these patients had a second colonoscopy at 3 years postoperatively. CEA levels were checked twice annually for 3 years according to surveillance guidelines in 56 (45.1%) only. However, Abdomen/pelvis CT scan was obtained in 90 (72.58%) patients at 1 year post-operatively.

## Discussion

We conducted a retrospective review aiming to assess the adherence rate to post-resection surveillance guidelines of CRC cases in two urban tertiary teaching medical centers and to evaluate the impact of demographics on strict compliance with guidelines. Our hypothesis was that the rate of adherence to guidelines was negatively influenced by certain patient demographics. In this study, we found (a) A low overall adherence rate to strict surveillance guidelines in patients who underwent curative resection of colorectal cancer, (b) this low adherence rate was not significantly impacted by insurance status, race, or gender, and (c) Our study also shows that these demographic features had no significant impact on the stage of cancer at the time of diagnosis.

After curative resection, recurrent disease arises in 30%-50% of cases, with the highest rate occurring within the first two years^9^. The yearly incidence of recurrent disease after curative resection is approximately 9.9% at 1 year, 26.2% at 3 years, and 31.5% at 5 years^9^. Hence, the guidelines advocate for an intensive follow-up program after primary surgical CRC treatment in the initial three-year period. The USMSTF has established surveillance guidelines after curative resection of colorectal cancer^10,11^.

Post-treatment surveillance is an essential component of the CRC survivorship plan, yet adherence to recommended guidelines among CRC survivors fell short of optimal levels. Challenges to surveillance include various factors such as limited access to healthcare, financial and psychological barriers, patient comorbidities, awareness and education gaps, and communication issues among healthcare providers^12^.

In this study, only 38% of patients received strict surveillance measures as outlined by USMSTF. Similarly, low rate of adherence to these guidelines has been found by Cooper *et al*. in a study of 9,426 patients who reported only 17.1% had appropriate surveillance^13^. In another study by Sisler *et al*., just 12.3% out of 250 patients received the recommended surveillance measures according to guidelines^14^. These low rates could be influenced by various barriers that could be related to the patient or the provider^15^. Patients undergoing uncomfortable bowel preparation for colonoscopy, and a lack of a designated responsible provider for conducting surveillance investigations among multiple healthcare providers were some of the identified factors impacting CRC survivor care^16^. Interventions to address and eliminate these and other surveillance barriers should be implemented to improve compliance. A comprehensive approach including patient education about its importance, implementation of patient support programs, addressing logistical barriers, training healthcare providers about guidelines and follow-up procedures, promoting clear communication among providers, and defining their roles in care plans^9^ are key to enhancing adherence for CRC survivors.

An important finding in our study is the lack of significant influence of demographic factors namely the type of insurance, race, patient’s gender, or the stage of cancer at the time of diagnosis on the post-resection surveillance in our cohort. These findings align with the previous results of Mohy-ud-din N *et al*., who found in a cohort of 80 patients that factors like age, gender, race, stage at diagnosis, and insurance status did not predict adherence to surveillance colonoscopy guidelines^13^. According to a study by Cooper *et al*., which involved 9,426 patients from the Medicare cancer database, adherence to guidelines was found to be associated with younger individuals, the white race, advanced-stage cancers (cancers that have spread to adjacent organs or regional lymph nodes), and poorly differentiated tumors. The study also showed variations in adherence rates based on geographical differences, which suggested that healthcare system and provider factors might have influenced the findings. Our study, on the other hand, included patients with varying insurance statuses, from the same geographic region who received treatment in the same clinical tertiary center. This consistency in location and provider-related factors enabled us to better assess how patient-specific elements affect adherence to guidelines.

Predicting survival rate of CRC is significantly determined by the stage at diagnosis, with the 5_year relative survival rate varying from 91% for localized disease to 14% for advanced disease^1^. The stage of CRC might be affected by screening practice and its utilization by the community, recognition of symptoms at presentation, and biological tumor growth variability^1^. Despite certain demographics that may influence the CRC incidence, it did not show a predictive value for the stage of CRC at the time of diagnosis in our cohort who underwent curative resection. This could be attributed to the homogeneity of patient characteristics, consistency in local healthcare practice, and uniform receipt of surgical treatment across all demographic groups in the study.

While there is no particular targeted intervention aimed at enhancing adherence to the surveillance recommended by clinical practice societies, different interventions have been explored. For instance, sending a mail reminder to the primary physician^14^ and implementing nurse coordinator supervision^15^ both have been shown to significantly enhance the adherence rate. Conversely, employing paper-based education approaches to the patients and providers did not yield success^16^. Another proposal was using electronic reminders followed by phone calls from staff members to encourage compliance^13^, which appears promising but further clinical research is required to validate the theory. In the future, additional data such as distance from the hospital, and other socioeconomic factors may help improve insight into patient surveillance following colorectal cancer resection.

Our study has certain limitations. As a retrospective study, a portion of our dataset was excluded due to a lack of known outcomes for patients, typically due to undocumented follow-ups. The majority of our cohort were African Americans, and the predominant type of insurance was Medicare/Medicaid which may limit the generalizability of our findings. However, at the same time, it may have provided valuable insight into these specific groups of patients. In addition, although our sample size only included 124 patients, 57% comprised men, and 44% were women which is fairly representative of CRC within the general population. However, the small sample size may have contributed to the discrepancy between our findings and those of previous reports. Larger cohort studies may eliminate this discrepancy in future studies.

## Conclusion

The current adherence rate to strict surveillance guidelines of colorectal cancer patients after curative resections is unacceptably low regardless of the patient’s demographic factors. Recognizing and addressing barriers to surveillance are critical measures to improve compliance with guidelines.

## Data Availability

The data used to support this study are included in the article. Further inquiries can be directed to the corresponding authors.

